# SmartAlert: Integrating Machine Learning and Alert Triggers into Live Electronic Medical Record Systems, Targeting Low-Yield Inpatient Lab Tests

**DOI:** 10.64898/2026.04.29.26351965

**Authors:** Yixing Jiang, Stephen P. Ma, April S. Liang, Grace Kim, Aakash Acharya, Sreedevi Mony, Soumya Punnathanam, John Makeown, Jeya Jose, Lisa Shieh, Tho Pham, Andrew Y. Ng, Jonathan H. Chen

## Abstract

This study explores integrating machine learning into electronic medical record systems to predict stability of inpatient lab tests. A ‘smart alerts’ system was developed and tested at Stanford Hospital. The system identifies stable lab results, advising clinicians on test ordering. Live deployment showed desired precision at good recall in predicting test result stability, with suggestions for system optimization identified. This approach may significantly decrease low-yield testing and enhance personalized clinical decision-making.

## Introduction

Low-yield repetitive laboratory testing not only burdens patients but also inflates the cost of care. Our prior study demonstrated the feasibility of using machine learning to identify low-yield repetitive tests (1). The objective of this study is to determine the feasibility of deploying custom machine learning models in a live electronic medical record (EMR) environment to predict whether common inpatient laboratory test panel components are stable.

## Methods

In this study, we identify a cohort of hospitalized patients for whom a clinician signed a repeat order for a lab test in our target group. Our target group for lab utilization optimization consists of the most common repetitive lab tests at Stanford Hospital and include Complete Blood Count (CBC), Basic Metabolic Panel, Serum Magnesium and Serum Phosphorus. The SmartAlert system identifies instances where all major components of that ordered lab are anticipated to remain stable. In these cases, the system may prompt clinicians to consider canceling or reducing the frequency of repeated testing.

Figure 1 shows the architecture of the SmartAlert system developed, based on the DEPLOYR framework (2). The first two components (a, b) are implemented on Microsoft Azure platform, and the third component (c) is configured on the Epic system. Because the process of extracting patient features and performing inference results in a significant delay when performed in real-time, we opted to perform inference in batches and cache the results for later review. The BatchInference function triggers the Inference function for all eligible individual patients in selected nursing units every six hours. The Inference function takes in a patient identifier, extracts features using Fast Healthcare Interoperability Resources (FHIR) APIs, and runs the probabilistic regression models. The prediction results are then saved to flowsheets in the Epic system. When a clinician is about to sign a repetitive lab test order, a Best Practice Alert (BPA) is displayed based on the results in the flowsheet and suggests decreasing lab utilization if the results are predicted to be stable.

**Figure 1.**
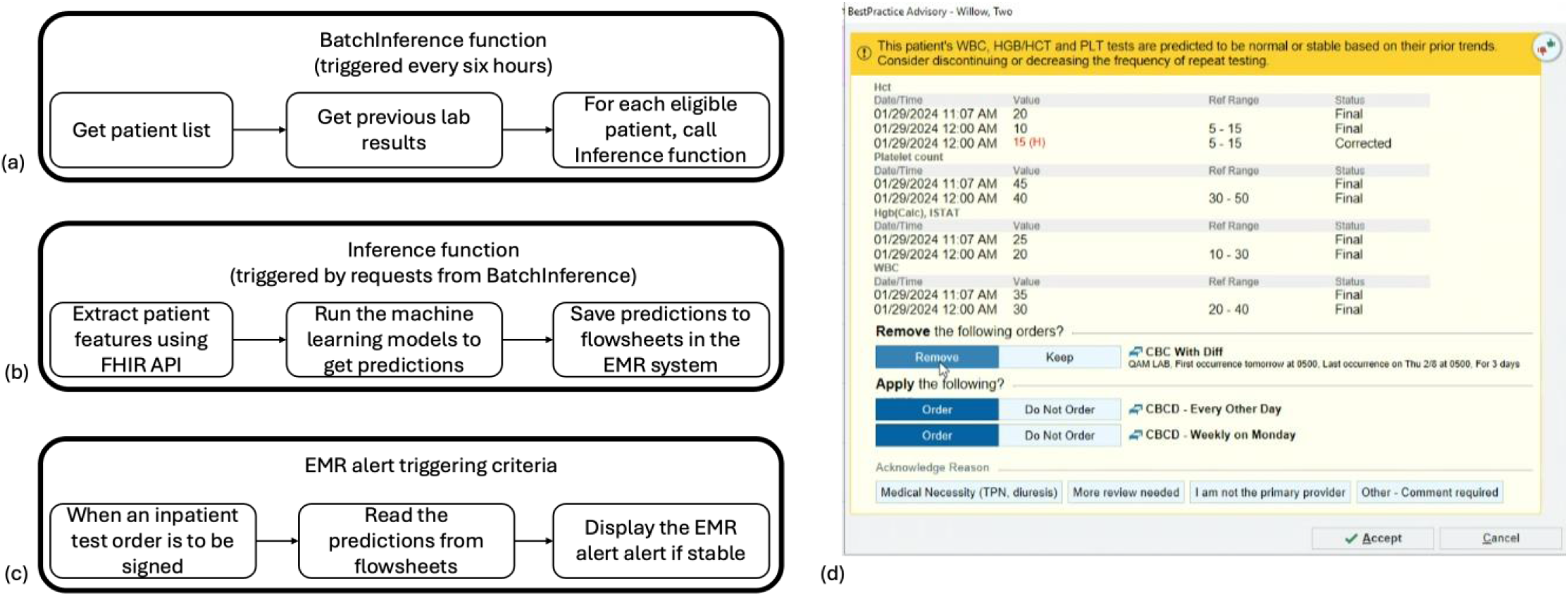
Diagram of overall system design. (a, b, c) shows the three back-end components; (d) shows one example screenshot of smart alerts.

In our prior work (1), we introduced the concept of a radius-based stability definition, where the repeat lab value is stable if it is within a certain range compared with its previous measure. The definition of stability is further optimized in this study based on clinician consensus review. We use asymmetric stability intervals due to differing clinical implications of changes, and the intervals are shown in Table 1. For example, a decrease in hemoglobin levels could signify blood loss, which is a clinically concerning event. Conversely, an increase in Hgb levels is less likely to indicate an acute issue, leading to a more permissive positive stability interval. Noting that the most common lab test orders are for panels of multiple components, we also utilized clinical consensus to identify which components are most important (e.g., Na, K, Ca, Cr for basic metabolic panel) and predict the stability of each component separately, while defining alert trigger logic to consider only cases where all of the key components are predicted to be stable.

**Table 1.** Recall at 90% precision in retrospective evaluation settings.

| Metric | WBC | HGB | PLT | NA | K | CR | CA | MG | PHOS |
| --- | --- | --- | --- | --- | --- | --- | --- | --- | --- |
| Recall at 90% PPV (%) | 92.7 | 21.1 | 96.4 | 41.9 | 100 | 100 | 100 | 100 | 22.5 |
| Stability interval | (-3.2, +2.4) | (-0.77, +2.00) | (-40.2, +90.0) | (-3.0, +3.4) | (-0.72, +0.94) | (-0.58, +0.26) | (-0.7, +0.86) | (-0.46, +0.78) | (-0.52, +1.26) |

To evaluate whether the SmartAlert system could work and maintain accurate stability predictions, we deployed the system silently from March 8, 2024 to March 14, 2024 and recorded stability predictions for 199 patients. We then compared the predicted stability statuses against the actual lab results that did occur in terms of common metrics including precision and recall.

## Results

Table 1 shows model evaluation results using a retrospective cohort during model development, and these results are reported as test performance in literature.

Our deployment in a live EMR system successfully triggered a full pipeline for live predictions of diagnostic test results stability. We compare predictions against actual measurements to assess model performance in a live setting, and the results are shown in Table 2. We targeted a threshold of 90% precision when training the model based on prior clinician feedback(3), which was maintained during prospective deployment. We also observe degradation on recall for some components like WBC and PLT while improvement on some other components like HGB, NA and PHOS. It is important to recognize these performance differences between retrospective and prospective settings and further highlight the value of live deployment.

**Table 2.** Model performances in prospective silent deployment evaluation settings.

| Metric | WBC | HGB | PLT | NA | K | CR | CA | MG | PHOS |
| --- | --- | --- | --- | --- | --- | --- | --- | --- | --- |
| Precision (%) | 88.1 | 95.4 | 93.0 | 88.7 | 92.8 | 96.9 | 93.2 | 95.7 | 88.0 |
| Recall (%) | 81.6 | 27.0 | 90.8 | 60.9 | 100 | 99.6 | 99.5 | 100 | 32.8 |

## Discussion and Conclusions

The integration of machine learning models into live EMR systems is now possible with API constructions such as the DEPLOYR framework, offering a promising avenue for enhancing clinical decision support in applications such as reducing low-yield laboratory testing. Our architecture allows for an end-to-end integrated smart alerts systems that spans triggering based on user EMR action to retrieving individual patient data, generating real-time inferences from custom machine learning models, and then writing the results back to the EMR to power personalized decision support logic. We were also able to further optimize this architecture through module I/O and startup processes to realize truly real-time delivery.

Our live silent deployment results show that our model maintains high levels of precision at different levels of recall for identifying low yield lab tests. These are at a level consistent with our qualitative studies with clinical focus groups on what level of certainty is sufficient to accept decision support guidance.

In conclusion, our findings illustrate that a significant proportion of inpatient labs can be predicted as stable with high accuracy using EMR data and custom machine learning models in a prospective, integrated setting. This capability presents a substantial opportunity to reduce low yield testing and can be extended to other personalized decision support applications.

## Data Availability

All data produced in the present work are contained in the manuscript

